# Perceptions of COVID-19 Vaccination During Pregnancy in a Cohort of US Adults (CHASING COVID) in 2023

**DOI:** 10.64898/2025.12.15.25342316

**Authors:** Rachael Piltch-Loeb, Subha Balasubramanian, McKaylee Robertson, Chloe Teasdale, Sasha Fleary, Josefina Nuñez Sahr, Denis Nash, Kate Penrose, Bai Xi Jasmine Chan, Angela Parcesepe

## Abstract

**Objective:** Pregnant women are at higher risk of severe COVID-19, and vaccination significantly reduces the risk of severe infection. Despite its benefits, only 13% of pregnant women in the U.S. had received the updated 2024–25 vaccine by December 2024, with uptake varying across sociodemographic groups. This study examines perceptions on COVID-19 vaccination during pregnancy among U.S. adults enrolled in the Chasing Covid Cohort, analyzing responses across 16 surveys between March 2020 and December 2023 (N=4488).

**Methods:** Key variables included sociodemographic characteristics, susceptibility to severe COVID-19 disease, perceived worry about COVID-19, individual and household vaccination status, symptoms of anxiety and depression, trusted information sources, and having a regular healthcare provider. Perceptions of vaccine safety and efficacy during pregnancy were measured using five Likert-scale statements, categorized into agreement, uncertainty, and disagreement. Exploratory factor analysis identified two constructs—safety and efficacy—which were analyzed in relation to participant characteristics using bivariate analysis and chi-square tests, and multivariable robust Poisson regression models.

**Results:** Among all respondents and women of reproductive age, less than half (40%) perceived the COVID-19 vaccine as safe during pregnancy, and just over half recognized its efficacy. Individuals with a personal physician and those who trusted public health institutions or healthcare providers were more inclined to agree with the vaccine’s safety and efficacy.

**Conclusions:** These findings highlight the influence of demographic factors on vaccine perceptions, the potential impact of social networks during pregnancy, and the critical role of trust in public health institutions in promoting vaccine uptake.

## 1. Background

Pregnant women and birthing people (“pregnant women”) are at higher risk of severe COVID-19[1,2]. Pregnant women with COVID-19 were more likely to be admitted to an intensive care unit (ICU), require invasive ventilation, require extracorporeal membrane oxygenation, and die when compared to nonpregnant women of reproductive age with COVID-19, according to the United States Centers for Disease Control and Prevention (CDC) COVID-19 surveillance system^1^. SARS-CoV-2 infection during pregnancy may also be associated with several adverse birth outcomes including preeclampsia, preterm birth, and stillbirth, especially among those with severe COVID-19 disease[2].

COVID-19 vaccination significantly reduces the risk of severe SARS-CoV-2 infection in the general population and during pregnancy[3]. Despite being largely excluded from the initial clinical trials, pregnant women have since been included in routine roll-out and follow-up studies focusing on the safety and efficacy of COVID-19 vaccination[4]. A 2021 cohort study of over 10,000 vaccinated pregnant women found the vaccine 89% effective preventing COVID-19-related hospitalization[5]. A 2022 global review found no major mRNA vaccine adverse events among pregnant women and confirmed their effectiveness preventing severe disease[3]. Additionally, maternal mRNA COVID-19 vaccination during pregnancy was associated with lower risks of severe neonatal morbidity, death, and ICU admission and no increase in neonatal readmission or hospital admission up to age 6 months[6]. Until infants are age-eligible for vaccination (six months), maternal vaccination provides passive protection against symptomatic infection[7].

The CDC and the American College of Obstetrics and Gynecology (ACOG) recommend COVID-19 vaccination during pregnancy [8,9]. However, as of December 2024, only 13% of pregnant women received an updated 2024-25 COVID-19 vaccine[10]. Uptake varies significantly by race/ethnicity - highest amongst non-Hispanic Asian (22%) and non-Hispanic White women (18%), with lower uptake observed among Hispanic (8%) and Black (6%) women[10], age (lower in younger women)[11], and education (highest with college education), urban residence and higher income [12,11]. Access to healthcare, including insurance, a primary care provider, or vaccination counseling from a healthcare provider, has been associated with higher odds of COVID-19 vaccination during pregnancy[13].

Despite ongoing efforts, significant gaps remain in understanding COVID-19 vaccine hesitancy during pregnancy. Addressing these gaps is crucial given the heightened risk of severe complications from COVID-19 for pregnant women and evolving vaccine recommendations. To gain a broader understanding of attitudes toward COVID-19 vaccination during pregnancy, we examined acceptability for pregnant women and its relationship with sociodemographics, healthcare experiences, risk perception, and trust in public health institutions in a cohort of US adults >=18 years and among women of reproductive age (18-49 years). Focusing on women of reproductive age highlights unique immunization concerns during/after pregnancy, while including all adults allows exploration of generational implications on vaccine attitudes. By deconstructing these diverse perspectives, this study hopes to provide significant insights for developing effective nuanced health communication that connects with a wide audience and addresses specific concerns of key subpopulations.

## 2. Methods

### 2.1 Study Population

The Communities, Households, and SARS-CoV-2 Epidemiology (CHASING) COVID Cohort study is a national prospective study initiated in March 2020, during the onset of the COVID-19 pandemic in the United States. We employed internet-based recruitment strategies to assemble a geographically and socio-demographically diverse cohort of participants aged ≥18 residing in the U.S. or its territories. Follow-up surveys were conducted approximately quarterly from March 2020 through December 2023. Further details on recruitment and procedures are available elsewhere[14]. Recruitment of the original study was completed between 28/03/2020 and 21/08/2020.This study was approved by the Institutional Review Boards of the City University of New York (CUNY) (New York, NY, USA) (protocol 2020-0256).

#### COVID-19 Vaccine Safety and Efficacy Perceptions During Pregnancy

We assessed perceptions of the COVID-19 vaccine’s safety and efficacy during pregnancy in December 2023 using responses to five specific statements: a) Not enough is known about the long-term side effects of receiving the COVID-19 vaccine during pregnancy; b) There is not enough research to support getting the COVID-19 vaccine during pregnancy; c) The COVID-19 vaccine is safe to receive during pregnancy; d) The COVID-19 vaccine reduces the risk of severe disease from COVID-19 during pregnancy; e) Receiving the COVID-19 vaccine while pregnant helps protect infants until they are old enough to be vaccinated.

Data were collected using a 5-point Likert scale, which was collapsed into three categories for analysis: “Agreement” (“strongly agree” or “agree”), “Uncertainty” (“neither agree nor disagree”), and “Disagreement” (“strongly disagree” or “disagree”). For two statements, responses were reverse-coded to align with the direction of the other statements.

### 2.2 Participant Characteristics

Age, gender, race/ethnicity, education, and household annual income were collected at cohort enrollment. Participants were classified as pregnant or recently pregnant if they reported being pregnant at any time between March 2020 and December 2023. Both cisgender women and non-binary individuals reporting a pregnancy were included. No transgender men reported a pregnancy. We identified participants with any children <18 years in the household as of October 2023.

#### a) Susceptibility to severe COVID-19

Susceptibility to severe COVID-19 disease was assessed at cohort enrollment using a composite variable based on factors the CDC identified as increasing risk for COVID-19 complications given SARS-CoV-2 infection[15]. These included age ≥60 years, self-report of chronic lung disease (CLD) or chronic obstructive pulmonary disease (COPD), current asthma, diabetes, serious heart conditions (including heart attack, high blood pressure, angina), kidney disease, immunocompromised status, HIV, or daily smoking. Each factor was coded as present (1) or absent (0), and a cumulative score was calculated by summing these binary variables, resulting in a score from 0-9. This score was then dichotomized based on the median as: more (score >1) or less (score ≤ 1) susceptible to severe COVID-19.

#### b) Vaccination status - individual and household

In over 16 rounds of follow-up surveys, participants were asked to report their personal and household members’ COVID-19 vaccination status. Participants were considered vaccinated if they reported completing their primary vaccination series (2 doses of a two-dose vaccine [e.g., mRNA vaccines] or 1 dose of a single-dose vaccine [e.g., J&J]). Participants were considered boosted in 2023 if they reported receiving a booster dose in 2023.

As of October 2023, if at least one COVID-19 vaccine-eligible person, including the enrolled participant, in a household, was COVID-19 vaccinated, the household was considered vaccinated.

#### c) Trusted information sources

In October 2023, participants were asked to identify various entities, groups, or individuals they trusted to provide reliable information about the COVID-19 vaccine (see Appendix). Trust in public health institutions was defined as endorsing trust in any of the following national or international public health agencies: CDC, the World Health Organization (WHO), or the Food and Drug Administration (FDA). Participants’ trust in healthcare providers was defined as endorsing personal physicians or other healthcare professionals as providers of reliable information about the COVID-19 vaccine.

#### d) Regular healthcare provider

Participants were categorized as having a personal doctor if they indicated they had one person they think of as their personal doctor or healthcare provider, as assessed in October 2023.

#### e) Symptoms of anxiety and depression

In December 2023, participants completed the seven-item Generalized Anxiety Disorder (GAD-7) scale and the eight-item Patient Health Questionnaire (PHQ-8). Those who scored ≥10 on the GAD-7 were classified as having moderate to severe anxiety symptoms, and those who scored ≥10 on the PHQ-8 were classified as having moderate to severe depression symptoms [16, 17].

#### f) Perceived worry

Assessed in December 2023, participants were coded as “worried” if they responded “somewhat worried” or “very worried” to either of the following questions: How worried are you about getting sick from COVID-19?; How worried are you about the long-term effects of COVID-19 infection(s)?

### 2.3 Analytic Approach

For this analysis, we included CHASING COVID cohort participants who completed enrollment surveys between March and July 2020, and who completed the December 2023 survey, which included questions about COVID-19 vaccination concerns during pregnancy. The analysis was conducted first among all adults and then only among women of reproductive age (18-49 years at enrollment). Individuals who did not answer all vaccine perception questions were excluded from the analysis. We calculated the proportion of respondents who agreed with each COVID-19 vaccine perception statement, indicating confidence in the vaccine.

#### Factor Analysis

We conducted an EFA to identify underlying constructs related to COVID-19 vaccine perceptions during pregnancy. The iterated principal axis factoring method with varimax rotation was used to extract two distinct factors: safety and efficacy perceptions. These categories were then used to classify participants according to their perceptions of safety and efficacy. Bivariate analyses examined associations between each identified factor and participant characteristics. Chi-square tests were utilized to explore the relationships between these characteristics, and the factors identified in the EFA.

#### Multivariable regression models

We used multivariable robust Poisson regression [18] to identify participant characteristics correlated with agreement that COVID-19 vaccination during pregnancy was safe or effective. We ran a separate model for each construct (vaccine safety and vaccine efficacy) and for two different comparisons: 1. “any agreement” versus “some disagreement,” excluding those categorized as uncertain, and 2. “any agreement” versus “uncertainty,” excluding those categorized as having some disagreement. We ran models among all adults and among women of reproductive age. This resulted in eight models across outcome, comparison, and population combinations. For each predictive model, we included age, gender, education, race/ethnicity, moderate-to-severe anxiety symptoms, moderate-to-severe depression symptoms, having a personal doctor, susceptibility to severe COVID-19, trust in public health institutions, and trust in healthcare providers. The model among all adults also included gender. All analyses were performed using SAS 9.4 (Cary, NC, USA).

## 3. Results

Of the 4537 participants from the CHASING COVID Cohort who completed the December 2023 survey, 4488 (99%) provided complete responses to the vaccine safety and efficacy statements during pregnancy questions. Table 1 characterizes the study participants. Of all participants, 36% (N=1,611) were women of reproductive age and, of those, 13% were pregnant or had been recently pregnant. Table 2 and Table 3 present summary statistics of COVID-19 vaccine perceptions during pregnancy within the study cohort among all adults and women of reproductive age, respectively. Among all adults, the proportion of participants uncertain about a statement ranged from 38% to 53%. Notably, more than half of the participants, 53% (N=2384), disagreed with the statement, “The COVID-19 vaccine reduces the risk of severe disease from COVID-19 during pregnancy”. Among women of reproductive age, 55% (N=879) disagreed with the statement “The COVID-19 vaccine reduces the risk of severe disease from COVID-19 during pregnancy”.

**Table 1.** Participant characteristics.

| Characteristics | All adults | Women of reproductive age<sup>a</sup> |
| --- | --- | --- |
| **Total, N** | **4488** | **1611** |
| **Age category, N (col %)** |  |  |
| Age category: 18-29 | 985 (21.9%) | 595 (36.9%) |
| Age category: 30-39 | 1299 (28.9%) | 606 (37.6%) |
| Age category: 40-49 | 832 (18.5%) | 410 (25.5%) |
| Age category: 50-64 | 600 (13.4%) | NA |
| Age category: 65+ | 772 (17.2%) | NA |
| **Gender, N (col %)** |  |  |
| Gender - Female/Non-binary<sup>a</sup> | 2510 (55.9%) | 1611 (100%) |
| Gender - Male | 1978 (44.1%) | NA |
| **Race/Ethnicity, N (col %)** |  |  |
| Race/Ethnicity - NH White | 2796 (62.3%) | 856 (53.1%) |
| Race/Ethnicity - Hispanic | 749 (16.7%) | 342 (21.2%) |
| Race/Ethnicity - NH Asian/PI/Other | 456 (10.2%) | 211 (13.1%) |
| Race/Ethnicity - NH Black | 487 (10.9%) | 202 (12.5%) |
| **Education, N (col %)** |  |  |
| Education - <high school/high school | 507 (11.3%) | 241 (15%) |
| Education - Some college | 1176 (26.2%) | 468 (29.1%) |
| Education - College graduate | 2805 (62.5%) | 902 (56%) |
| **Household annual income, N (col %)** |  |  |
| Income - <\$50k/Unknown | 1834 (40.9%) | 748 (46.4%) |
| Income - \$50k-\$100k | 1424 (31.8%) | 477 (29.6%) |
| Income - >\$100k | 1227 (27.4%) | 386 (24%) |
| **Any <18 y in household, N (col %)** |  |  |
| Any <18y kid in household - No | 2543 (56.7%) | 722 (44.8%) |
| Any <18y kid in household - Yes | 1945 (43.3%) | 889 (55.2%) |
| **Pregnant, N (col %)** |  |  |
| Ever pregnant | 210 (4.7%) | 209 (13%) |
| Never pregnant | 4278 (95.3%) | 1402 (87%) |
| **Moderate to severe symptoms of anxiety as of December 2023, N (col %)** |  |  |
| Moderate to severe symptoms of anxiety - No | 3839 (85.5%) | 1288 (80%) |
| Moderate to severe symptoms of anxiety - Yes | 649 (14.5%) | 323 (20.1%) |
| **Depression status (PHQ) as of December 2023, N (col %)** |  |  |
| Depression status (PHQ) - No | 3682 (82%) | 1230 (76.4%) |
| Depression status (PHQ) - Yes | 806 (18%) | 381 (23.7%) |
| **Has personal doctor as of December 2023, N (col %)** |  |  |
| Has personal doctor - No | 1050 (24.5%) | 483 (31.3%) |
| Has personal doctor - Yes | 3237 (75.5%) | 1061 (68.7%) |
| **Susceptible to severe COVID-19 (if infected) at baseline, N (col %)** |  |  |
| Susceptible to severe COVID-19 (if infected) - No | 3527 (78.6%) | 1447 (89.8%) |
| Susceptible to severe COVID-19 (if infected) - Yes | 961 (21.4%) | 164 (10.2%) |
| **Vaccination status as of December 2023, N (col %)** |  |  |
| Vaccination status - No | 63 (1.5%) | 34 (2.5%) |
| Vaccination status - Yes | 4030 (98.5%) | 1354 (97.6%) |
| **Received Booster dose in 2023 N (col %)** |  |  |
| Boosted in 2023 - No | 2653 (59.1%) | 1137 (70.6%) |
| Boosted in 2023 - Yes | 1835 (40.9%) | 474 (29.4%) |
| **Household members vaccinated as of October 2023, N (col %)** |  |  |
| Household members vaccinated - No | 504 (11.8%) | 275 (17.8%) |
| Household members vaccinated - Yes | 3774 (88.2%) | 1266 (82.2%) |
| **Trust in government sources as of October 2023, N (col %)** |  |  |
| Trust in government information - No | 926 (21.7%) | 377 (24.5%) |
| Trust in government information - Yes | 3339 (78.3%) | 1160 (75.5%) |
| **Trust in healthcare providers as of October 2023, N (col %)** |  |  |
| Trust in healthcare providers - No | 1267 (29.7%) | 581 (37.8%) |
| Trust in healthcare providers - Yes | 2998 (70.3%) | 956 (62.2%) |
| **Perceived worry, as of December 2023 N (col %)** |  |  |
| Perceived worry - No | 2372 (52.9%) | 762 (47.3%) |
| Perceived worry - Yes | 2116 (47.1%) | 849 (52.7%) |

<sup>a</sup>We classify all gender minorities (N = 129) with women due to small numbers and in recognition of the increased health risks in this population.

**Table 2.** Summary Statistics of COVID-19 Vaccine Perception Statements as reported by all adults >=18 years in the CHASING COVID Cohort (N=4488)

| N (row%) | Agreement<sup>b</sup> | Uncertainty<sup>c</sup> | Disagreement<sup>d</sup> |
| --- | --- | --- | --- |
| a) Enough is known about the long-term side effects of receiving the COVID-19 vaccine during pregnancy<sup>a</sup> | 1351 (30.1%) | 1699 (37.8%) | 1438 (32.0%) |
| b) There is enough research to support getting the COVID-19 vaccine during pregnancy<sup>a</sup> | 1047 (23.3%) | 1806 (40.2%) | 1635 (36.4%) |
| c) The COVID-19 vaccine is safe to receive during pregnancy | 468 (10.4%) | 2112 (47.0%) | 1908 (42.5%) |
| d) The COVID-19 vaccine reduces the risk of severe disease from COVID-19 during pregnancy | 322 (7.2%) | 1782 (39.7%) | 2384 (53.1%) |
| e) Receiving the COVID-19 vaccine while pregnant helps protect infants until they are old enough to be vaccinated | 401 (8.9%) | 2386 (53.2%) | 1701 (37.9%) |

<sup>a</sup>Statements have been reworded and reverse coded to ensure consistent interpretation across all statements.
<sup>b</sup>Participants who expressed agreement with at least one statement within the factor were categorized as agreeing with the belief that COVID-19 vaccination during pregnancy is safe and effective.
<sup>c</sup>Participants who expressed uncertainty for all statements within the factor were categorized as having uncertainty
<sup>d</sup>Participants who expressed uncertainty or disagreement for all statements within the factor, but not exclusively uncertainty, were categorized as having some disagreement.
Note: We classify all gender minorities (N = 129) with women due to small numbers and in recognition of the increased health risks in this population.

**Table 3:**
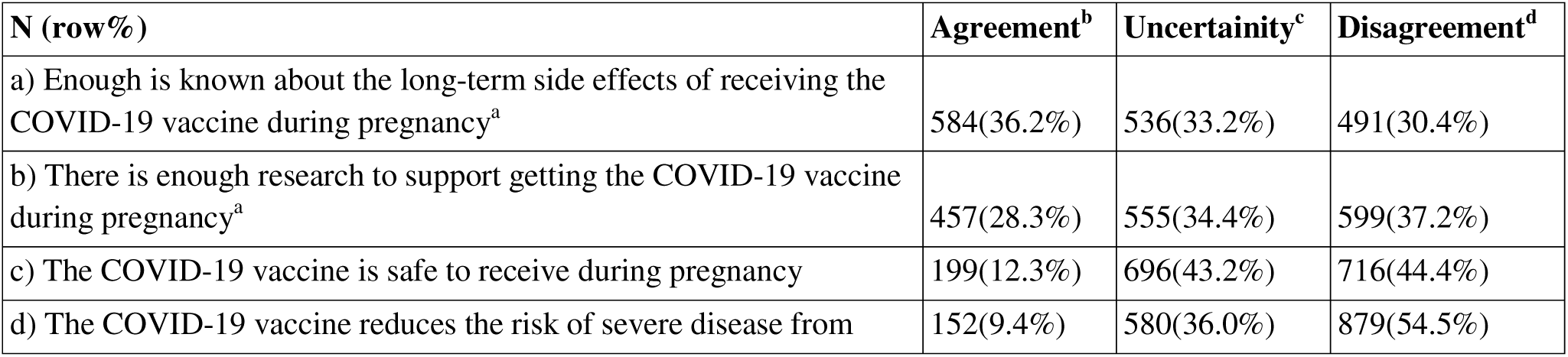
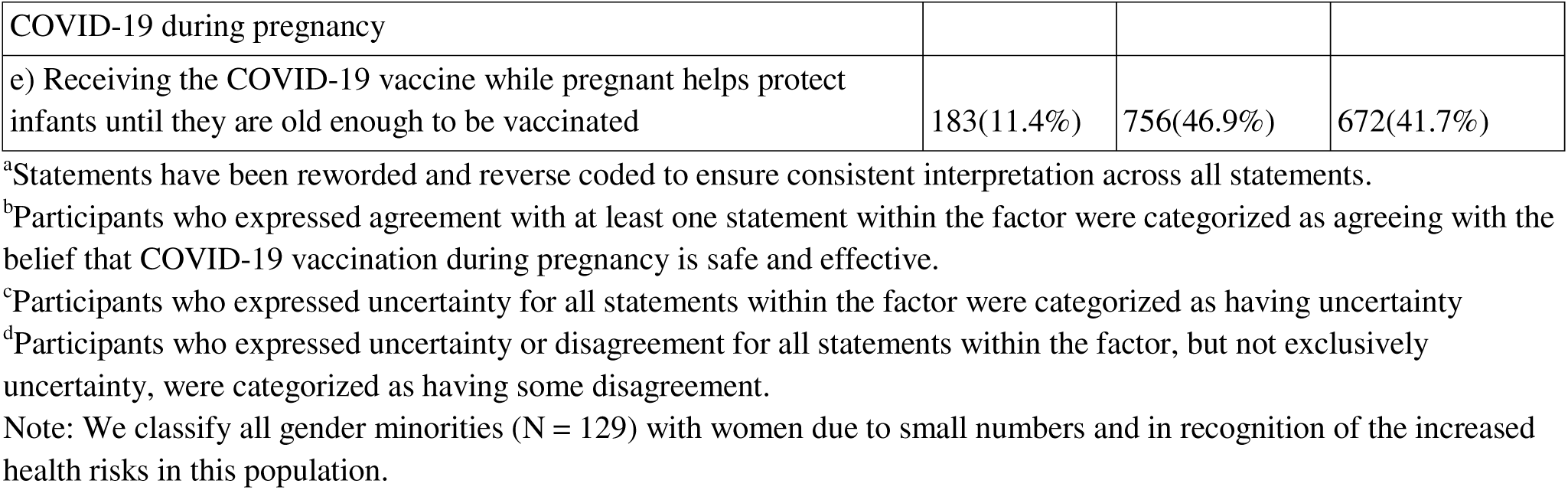
Summary Statistics of COVID-19 Vaccine Perception Statements as reported by women of reproductive age in the CHASING COVID Cohort (N=1611)

### Factor Analysis

As seen in Table 4, the EFA identified two factors related to any agreement with vaccine perceptions: safety and efficacy. For all adults, the items loaded significantly onto these two factors, with statements regarding the long-term side effects, safety, and sufficient research to support vaccination positively loading on the safety factor, while statements about the protection of infants and reduction of risk of severe disease positively loaded on the efficacy factor. The factor analysis for the subpopulation of women of reproductive age showed similar patterns. The reliability analysis conducted among all adults yielded a Cronbach’s alpha of 0.84 for safety perceptions and 0.85 for efficacy perceptions, indicating good internal consistency. The proportions of safety and efficacy perceptions were comparable between all adults and the subpopulation of women of reproductive age. Among both groups, more than half expressed any agreement with COVID-19 vaccine efficacy statements, with 58.2% of all adults and 59.7% of women of reproductive age reporting any agreement (Table 5).

**Table 4.**
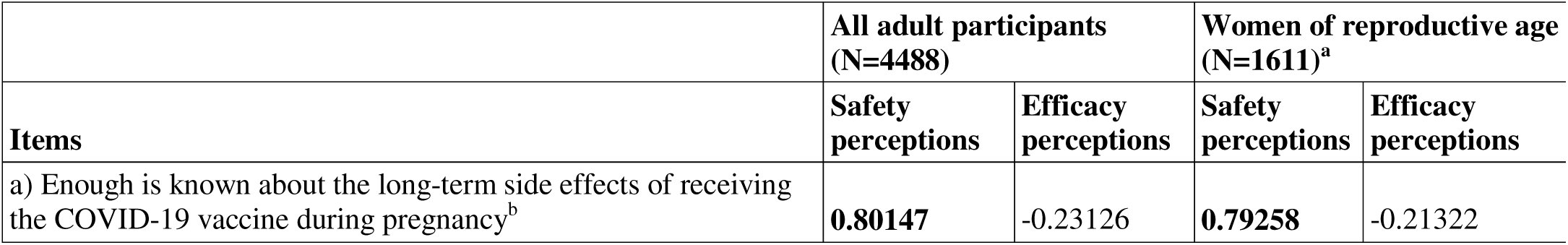
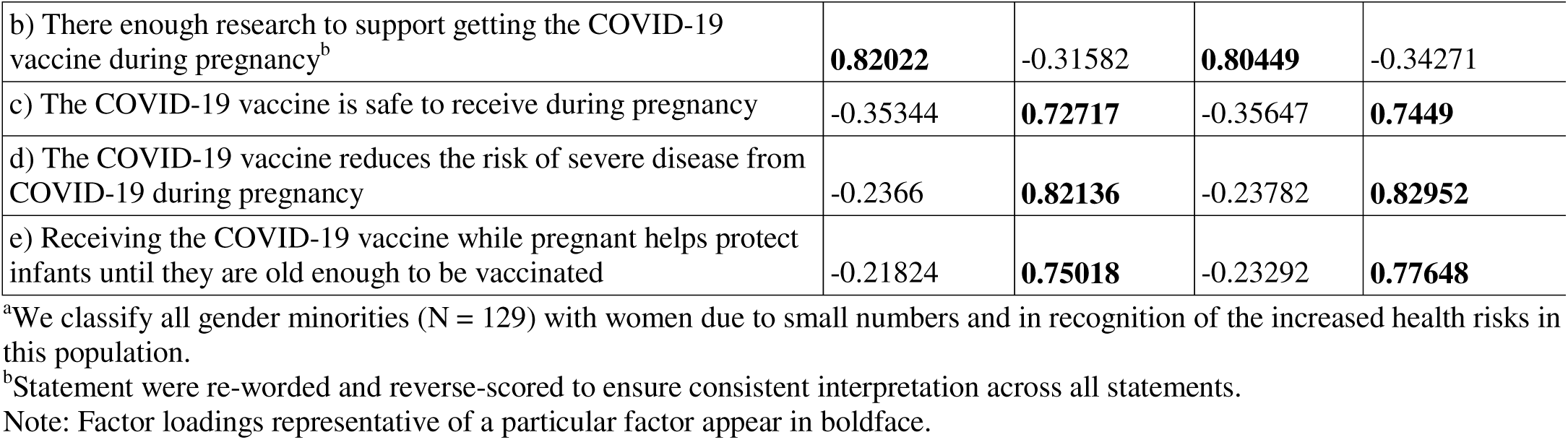
Vaccine perceptions during pregnancy scale items factor loading matrix (EFA)

|  | All adult participants<br>(N=4488) |  | Women of reproductive age<br>(N=1611)<sup>a</sup> |  |
| --- | --- | --- | --- | --- |
| Items | Safety<br>perceptions | Efficacy<br>perceptions | Safety<br>perceptions | Efficacy<br>perceptions |
| a) Enough is known about the long-term side effects of receiving the COVID-19 vaccine during pregnancy<sup>b</sup> | **0.80147** | -0.23126 | **0.79258** | -0.21322 |
| b) There enough research to support getting the COVID-19 vaccine during pregnancy<sup>b</sup> | **0.82022** | -0.31582 | **0.80449** | -0.34271 |
| c) The COVID-19 vaccine is safe to receive during pregnancy | -0.35344 | **0.72717** | -0.35647 | **0.7449** |
| d) The COVID-19 vaccine reduces the risk of severe disease from COVID-19 during pregnancy | -0.2366 | **0.82136** | -0.23782 | **0.82952** |
| e) Receiving the COVID-19 vaccine while pregnant helps protect infants until they are old enough to be vaccinated | -0.21824 | **0.75018** | -0.23292 | **0.77648** |

<sup>a</sup>We classify all gender minorities (N = 129) with women due to small numbers and in recognition of the increased health risks in this population.
<sup>b</sup>Statement were re-worded and reverse-scored to ensure consistent interpretation across all statements.
Note: Factor loadings representative of a particular factor appear in boldface.

**Table 5:** Alpha reliabilities and mean (SD) and median (IQR) COVID-19 perceptions during pregnancy by population.

|  | All adult participants (N=4488) |  |  | Women of reproductive age (N=1611)<sup>a</sup> |  |  | Cronbach's alpha |
| --- | --- | --- | --- | --- | --- | --- | --- |
|  | Any agreement<sup>b</sup> | Uncertainty<sup>c</sup> | Some disagreement<sup>d</sup> | Any agreement<sup>b</sup> | Uncertainty<sup>c</sup> | Some disagreement<sup>d</sup> |  |
| Safety perceptions | 1815 (40.4%) | 1367 (30.5%) | 1306 (29.1%) | 655 (40.7%) | 389 (24.1%) | 567 (35.2%) | 0.84 |
| Efficacy perceptions | 2611 (58.2%) | 1390 (30.9%) | 487 (10.9%) | 961 (59.7%) | 434 (26.9%) | 216 (13.4%) | 0.85 |

<sup>a</sup>We classify all gender minorities (N = 129) with women due to small numbers and in recognition of the increased health risks in this population.
<sup>b</sup>Participants who expressed agreement with at least one statement within the factor were categorized as agreeing with the belief that COVID-19 vaccination during pregnancy is safe and effective.
<sup>c</sup>Participants who expressed uncertainty for all statements within the factor were categorized as having uncertainty
<sup>d</sup>Participants who expressed uncertainty or disagreement for all statements within the factor, but not exclusively uncertainty, were categorized as having some disagreement.
Note: Statements a) and b) have been reworded and reverse coded to ensure consistent interpretation across all statements.

### Participant characteristics associated with vaccine safety and efficacy perceptions

Among all adults, 40.4% had any agreement, 30.5% were uncertain, and 29.1% had some disagreement with the statements regarding the safety of the COVID-19 vaccine during pregnancy; 58.2% had any agreement, 30.9% were uncertain, and 10.9% had some disagreement with the statements concerning the vaccine’s efficacy (Table 5).

Participants aged 30-39, non-Hispanic White individuals, and college graduates without children were more likely to have any agreement with statements about the safety and efficacy of the COVID-19 vaccine during pregnancy compared to those who expressed some disagreement (Table 6). Additionally, participants who did not report moderate to severe symptoms of anxiety or depression, and those who reported being fully vaccinated as of December 2023 or boosted in 2023, had a personal physician, and expressed trust in the public health institutions or healthcare providers were more likely to have any agreement with statements about the COVID-19 vaccine’s safety and efficacy during pregnancy than those who expressed some disagreement. Chi-square tests indicated that all characteristics were significantly associated with safety perceptions of the COVID-19 vaccine during pregnancy. However, moderate to severe symptoms of anxiety and depression were not significantly associated with efficacy perceptions of the vaccine during pregnancy (Table 6).

**Table 6.**
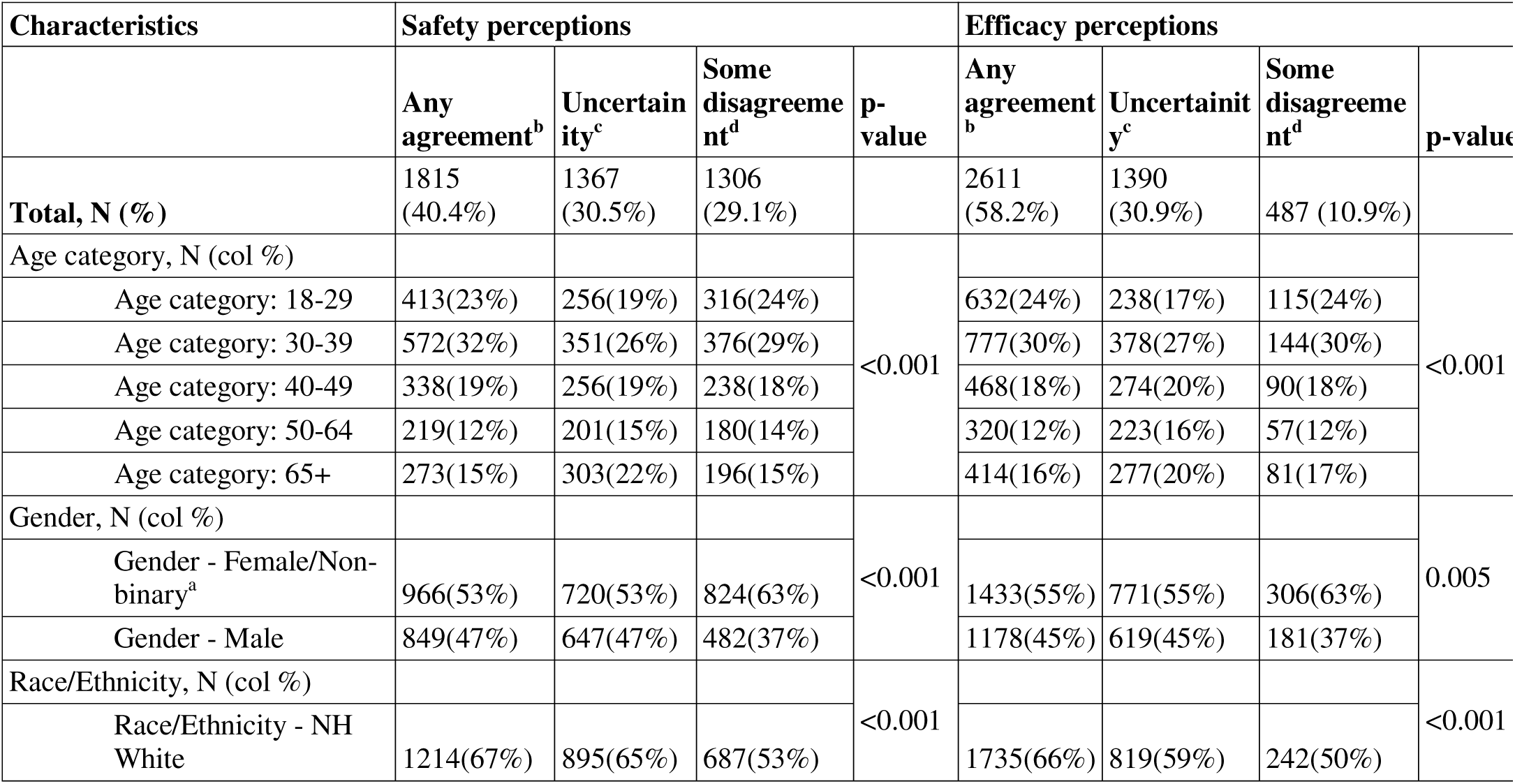
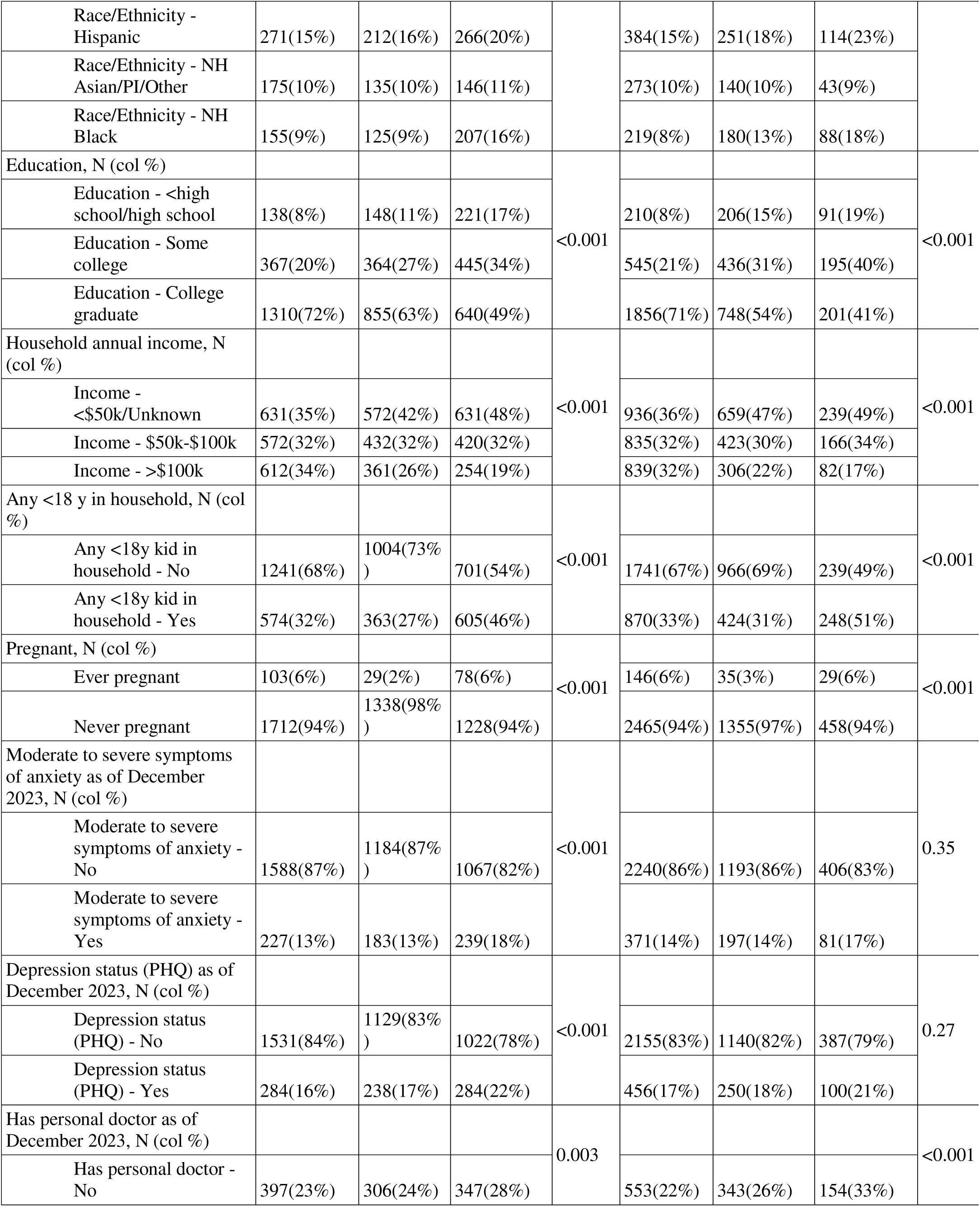
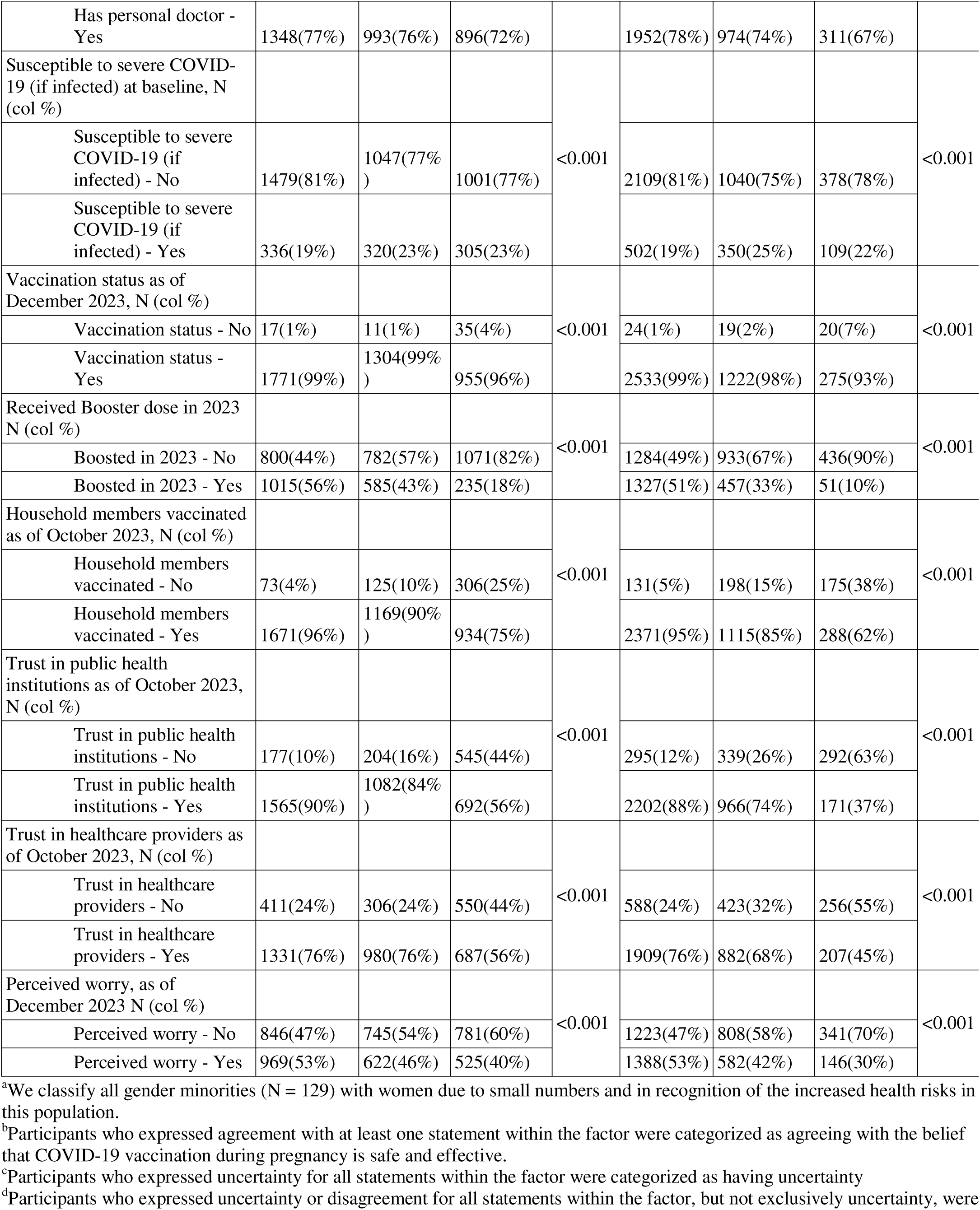
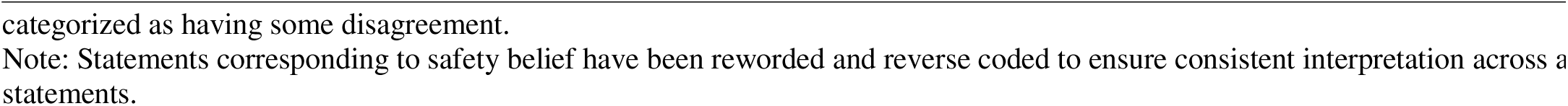
Participant characteristics by perception towards COVID vaccine concerns during pregnancy among all adult participants (N=4488)

While findings were similar among women of reproductive age, in that group, women aged 18-29 and those more worried about getting sick again from COVID-19 were more likely to have any agreement about the safety and efficacy statements of the COVID-19 vaccine during pregnancy (Table 7). Similar to all adults, moderate to severe symptoms of anxiety and depression were not significantly associated with perceptions of COVID-19 vaccine efficacy (Table 7).

**Table 7.**
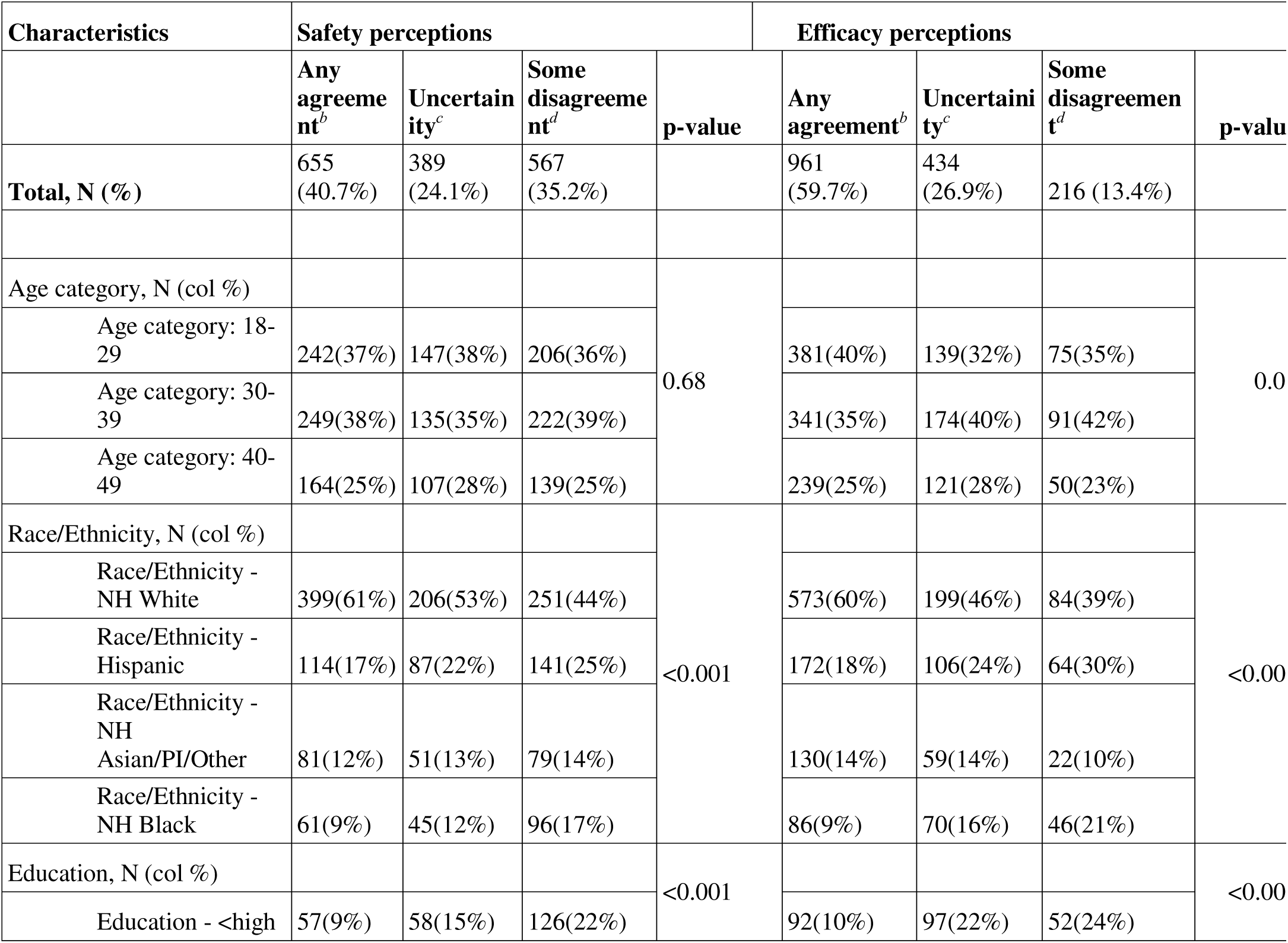
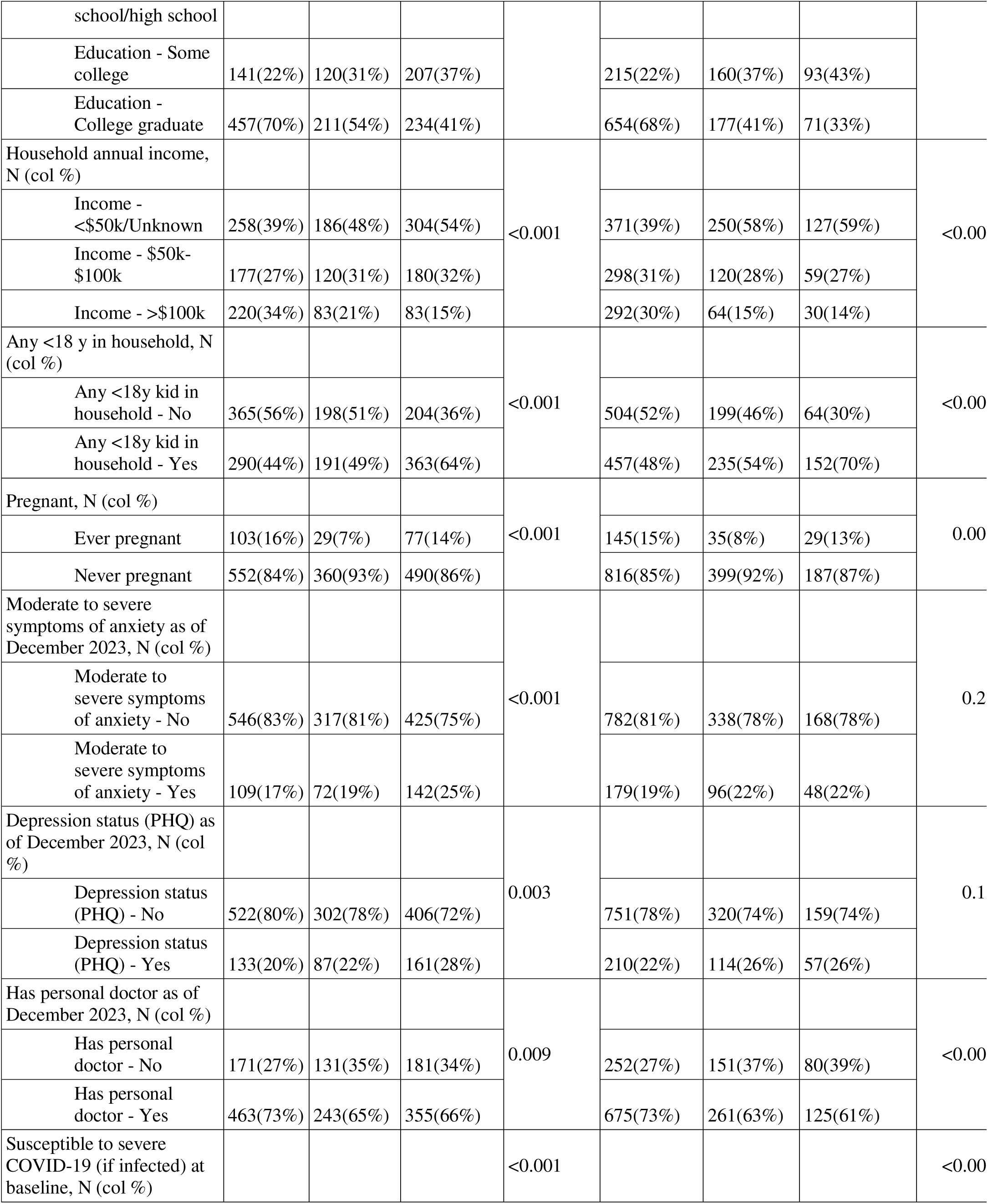
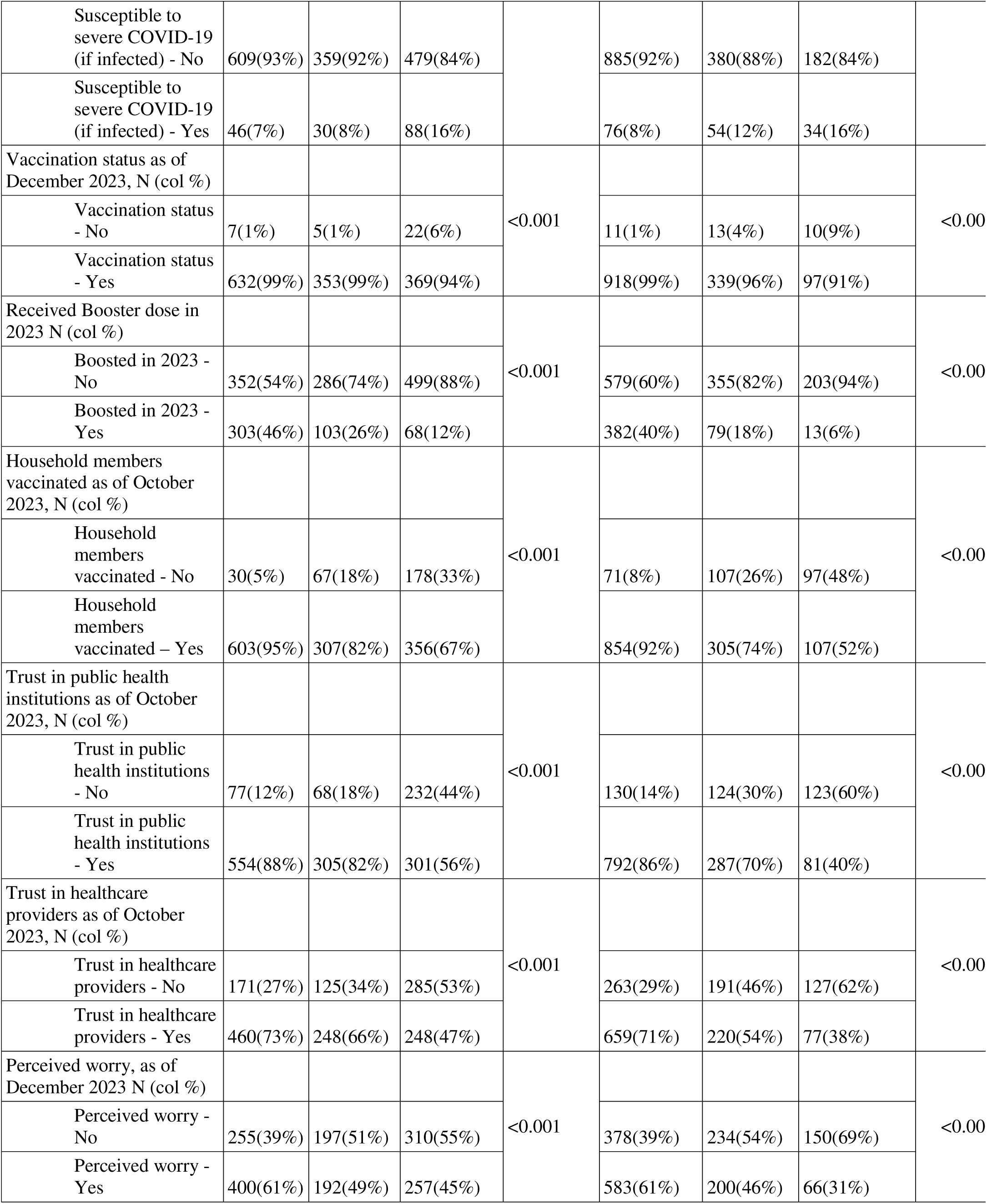
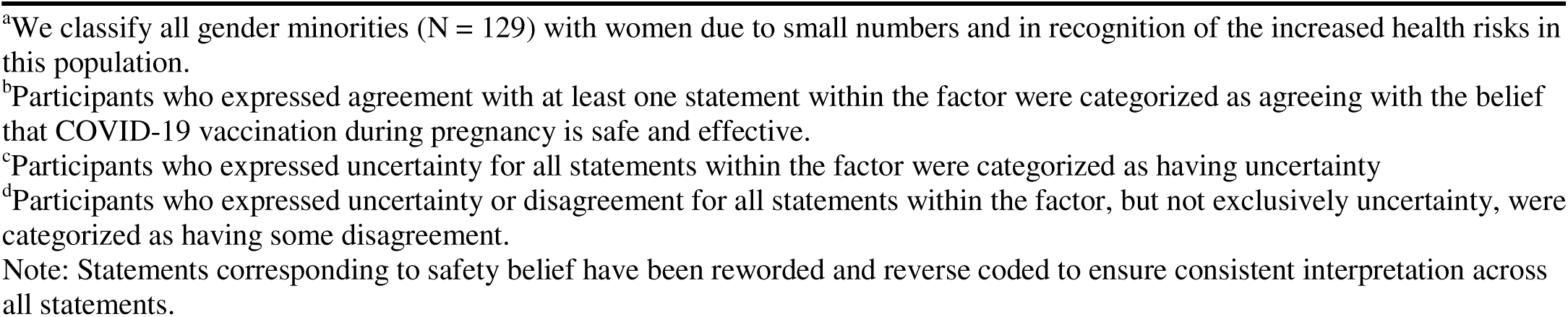
Participant characteristics by perception towards COVID vaccine concerns during pregnancy among women^a^ of reproductive age (N=1611)

### Multivariate Models

#### All adults

Table 8 presents unadjusted and adjusted prevalence ratios (aPRs) for any agreement vs. some disagreement with COVID-19 vaccine statements during pregnancy in the cohort (N=4,488). Participants who reported trusting public health institutions (aPR=2.48, 95% CI: 2.17–2.83) and trusting healthcare providers (aPR=1.23, 95% CI: 1.14–1.33) were more likely to have any agreement about statements related to the safety of COVID-19 vaccine during pregnancy compared to those without trust in information sources. Conversely, symptoms of anxiety (aPR=1.00, 95% CI: 0.89–1.13) and depression (aPR=0.96, 95% CI: 0.87–1.07) were not significantly associated with COVID-19 vaccine statements during pregnancy. Non-Hispanic Black participants were 19% less likely than their non-Hispanic White counterparts to have any agreement with statements related to the vaccine’s safety during pregnancy (aPR=0.81, 95% CI: 0.71–0.92).

**Table 8.** Prevalence Ratios and P-values for Any Agreement vs. Some Disagreement about COVID-19 Vaccine During Pregnancy in the CHASING COVID Cohort among all adult participants.

|  | Safety perceptions (N = 3,121) |  |  |  | Efficacy perceptions (N = 3,098) |  |  |  |
| --- | --- | --- | --- | --- | --- | --- | --- | --- |
|  | Prevalence Ratio | P-value | Adjusted Prevalence ratio<sup>a</sup> | P-value | Prevalence Ratio | P-value | Adjusted Prevalence ratio<sup>a</sup> | P-value |
| **Age** 18-29 vs 30-39 | 0.94<br>(0.87 - 1.02) | 0.13 | 1.03<br>(0.96 - 1.12) | 0.41 | 1.00<br>(0.96 - 1.05) | 0.89 | 1.05<br>(1.01 - 1.09) | 0.03 |
| **Age** 40-49 vs 30-39 | 0.97<br>(0.89 - 1.06) | 0.53 | 0.97<br>(0.89 - 1.05) | 0.41 | 0.99<br>(0.95 - 1.04) | 0.80 | 0.98<br>(0.94 - 1.02) | 0.43 |
| **Age** 50-64 vs 30-39 | 0.91<br>(0.82 - 1.01) | 0.07 | 0.89<br>(0.81 - 0.97) | 0.01 | 1.01<br>(0.96 - 1.06) | 0.81 | 0.98<br>(0.94 - 1.02) | 0.41 |
| **Age** 65+ vs 30-39 | 0.96<br>(0.88 - 1.06) | 0.45 | 0.98<br>(0.89 - 1.07) | 0.61 | 0.99<br>(0.95 - 1.04) | 0.72 | 0.97<br>(0.92 - 1.02) | 0.26 |
| **Gender** Male vs Female/Non-binary<sup>b</sup> | 1.18<br>(1.11 - 1.25) | <0.001 | 1.11<br>(1.05 - 1.17) | <0.001 | 1.05<br>(1.02 - 1.08) | 0.001 | 1.03<br>(1.00 - 1.06) | 0.02 |
| **Race/Ethnicity** Hispanic vs NH White | 0.79<br>(0.72 - 0.86) | <0.001 | 0.94<br>(0.86 - 1.02) | 0.13 | 0.88<br>(0.84 - 0.92) | <0.001 | 0.95<br>(0.90 - 0.99) | 0.03 |
| **Race/Ethnicity** NH Black vs NH White | 0.67<br>(0.59 - 0.76) | <0.001 | 0.81<br>(0.71 - 0.92) | <0.001 | 0.81<br>(0.76 - 0.87) | <0.001 | 0.91<br>(0.85 - 0.97) | 0.006 |
| **Race/Ethnicity** NH Asian/PI/Other vs NH White | 0.85<br>(0.77 - 0.95) | 0.003 | 0.89<br>(0.81 - 0.99) | 0.02 | 0.98<br>(0.94 - 1.03) | 0.51 | 1.00<br>(0.97 - 1.05) | 0.82 |
| **Income** \$50k-\$100k vs <\$50k/Unknown | 1.15<br>(1.07 - 1.25) | 0.003 | NA | | 1.05<br>(1.01 - 1.09) | 0.02 | NA | |
| **Income** >\$100k vs <\$50k/Unknown | 1.41<br>(1.32 - 1.52) | <0.001 | NA | | 1.14<br>(1.10 - 1.18) | <0.001 | NA | |
| **Education** <high school/high school vs College graduate | 0.57<br>(0.50 - 0.65) | <0.001 | 0.73<br>(0.64 - 0.84) | <0.001 | 0.77<br>(0.72 - 0.83) | <0.001 | 0.89<br>(0.82 - 0.95) | <0.001 |
| **Education** Some college vs College graduate | 0.67<br>(0.62 - 0.73) | <0.001 | 0.82<br>(0.76 - 0.89) | <0.001 | 0.82<br>(0.78 - 0.85) | <0.001 | 0.91<br>(0.87 - 0.95) | <0.001 |
| **Anxiety status** moderate/sev vs none to mild | 0.81<br>(0.74 - 0.90) | <0.001 | 1.00<br>(0.89 - 1.13) | 0.96 | 0.97<br>(0.93 - 1.02) | 0.19 | 1.06<br>(1.01 - 1.12) | 0.02 |
| **Depression status** moderate/sev vs none to mild | 0.83<br>(0.76 - 0.91) | <0.001 | 0.96<br>(0.87 - 1.07) | 0.49 | 0.97<br>(0.93 - 1.01) | 0.12 | 1.00<br>(0.95 - 1.05) | 0.89 |
| **Any <18y kid in household** Yes vs No | 0.76<br>(0.71 - 0.82) | <0.001 | NA |  | 0.89<br>(0.85 - 0.92) | <0.001 | NA |  |
| **Pregnancy Status**<br>Ever pregnant vs<br>Never pregnant | 0.98<br>(0.86 - 1.11) | 0.73 | NA |  | 0.99<br>(0.92 - 1.06) | 0.76 | NA |  |
| **Personal doctor** Yes<br>vs No | 1.13<br>(1.04 - 1.21) | 0.002 | 1.00<br>(0.94 - 1.08) | 0.90 | 1.10<br>(1.06 - 1.15) | <0.001 | 1.05<br>(1.01 - 1.09) | 0.01 |
| **Susceptibility to**<br>**severe COVID-19**<br>**(if infected)** at<br>baseline above vs<br>below | 0.88<br>(0.81 - 0.95) | 0.002 | 0.94<br>(0.86 - 1.02) | 0.14 | 0.97<br>(0.93 - 1.01) | 0.13 | 1.00<br>(0.96 - 1.05) | 0.96 |
| **Vaccination status**<br>Yes vs No | 1.99<br>(1.34 - 2.94) | 0.001 | NA |  | 1.65<br>(1.26 - 2.17) | <0.001 | NA |  |
| Boosted in 2023 Yes<br>vs No | 1.90<br>(1.79 - 2.01) | <0.001 | NA |  | 1.29<br>(1.25 - 1.33) | <0.001 | NA |  |
| **Household**<br>**members**<br>**vaccinated** Yes vs<br>No | 3.33<br>(2.70 - 4.10) | <0.001 | NA |  | 2.08<br>(1.83 - 2.37) | <0.001 | NA |  |
| **Trust** in public<br>health institutions<br>Yes vs No | 2.83<br>(2.48 - 3.22) | <0.001 | 2.48<br>(2.17 - 2.83) | <0.001 | 1.85<br>(1.70 - 2.00) | <0.001 | 1.73<br>(1.59 - 1.87) | <0.001 |
| **Trust** in healthcare<br>providers Yes vs No | 1.54<br>(1.42 - 1.67) | <0.001 | 1.23<br>(1.14 - 1.33) | <0.001 | 1.29<br>(1.24 - 1.36) | <0.001 | 1.13<br>(1.08 - 1.17) | <0.001 |
| **Perceived worry** as<br>of December 2023<br>Yes vs No | 1.20<br>(1.13 - 1.28) | <0.001 | NA |  | 1.14<br>(1.11 - 1.17) | <0.001 | NA |  |

<sup>a</sup>Fully adjusted model, including age, race/ethnicity, education, anxiety status, depression status, personal doctor, susceptibility to severe COVID-19, trust in public health institutions and trust in healthcare providers.
<sup>b</sup>We classify all gender minorities (N = 129) with women due to small numbers and in recognition of the increased health risks in this population.
Note: Statements corresponding to safety belief have been re-worded and reverse coded to ensure consistent interpretation across all statements.

Regarding associations with vaccine efficacy, trust in public health institutions (aPR=1.73, 95% CI: 1.59–1.87), trust in healthcare providers (aPR=1.13, 95% CI: 1.08–1.17), and having a personal doctor (aPR=1.05, 95% CI: 1.01–1.09) were statistically significant. Non-Hispanic Black participants were 9% less likely to have any agreement with statements related to the vaccine’s efficacy (aPR=0.91, 95% CI: 0.85-0.97) during pregnancy compared to their non-Hispanic White counterparts.

#### Women of reproductive age

Table 9 presents unadjusted and adjusted prevalence ratios (aPRs) for any agreement vs. some disagreement with COVID-19 vaccine statements during pregnancy among women of reproductive age in the cohort (N=1,611). Participants who trusted healthcare providers (aPR=1.34, 95% CI: 1.18–1.52) were 34% more likely to agree with statements related to the vaccine’s safety during pregnancy compared to those who did not trust healthcare providers. Similarly, participants who trusted public health institutions (aPR=2.15, 95% CI: 1.76–2.63) were more than twice as likely to agree with these statements compared to those who did not trust public health institutions. However, while the relationship between having a personal physician (aPR=1.03, 95% CI: 0.92–1.15) and perceptions about the vaccine’s safety suggests a positive association, it is not statistically significant at the 0.05 level. Women more susceptible to severe COVID-19 if infected with SARS-CoV-2 were 25% less likely to have any agreement with statements about the vaccine’s safety during pregnancy than those less susceptible (aPR=0.75, 95% CI: 0.59–0.95).

**Table 9.**
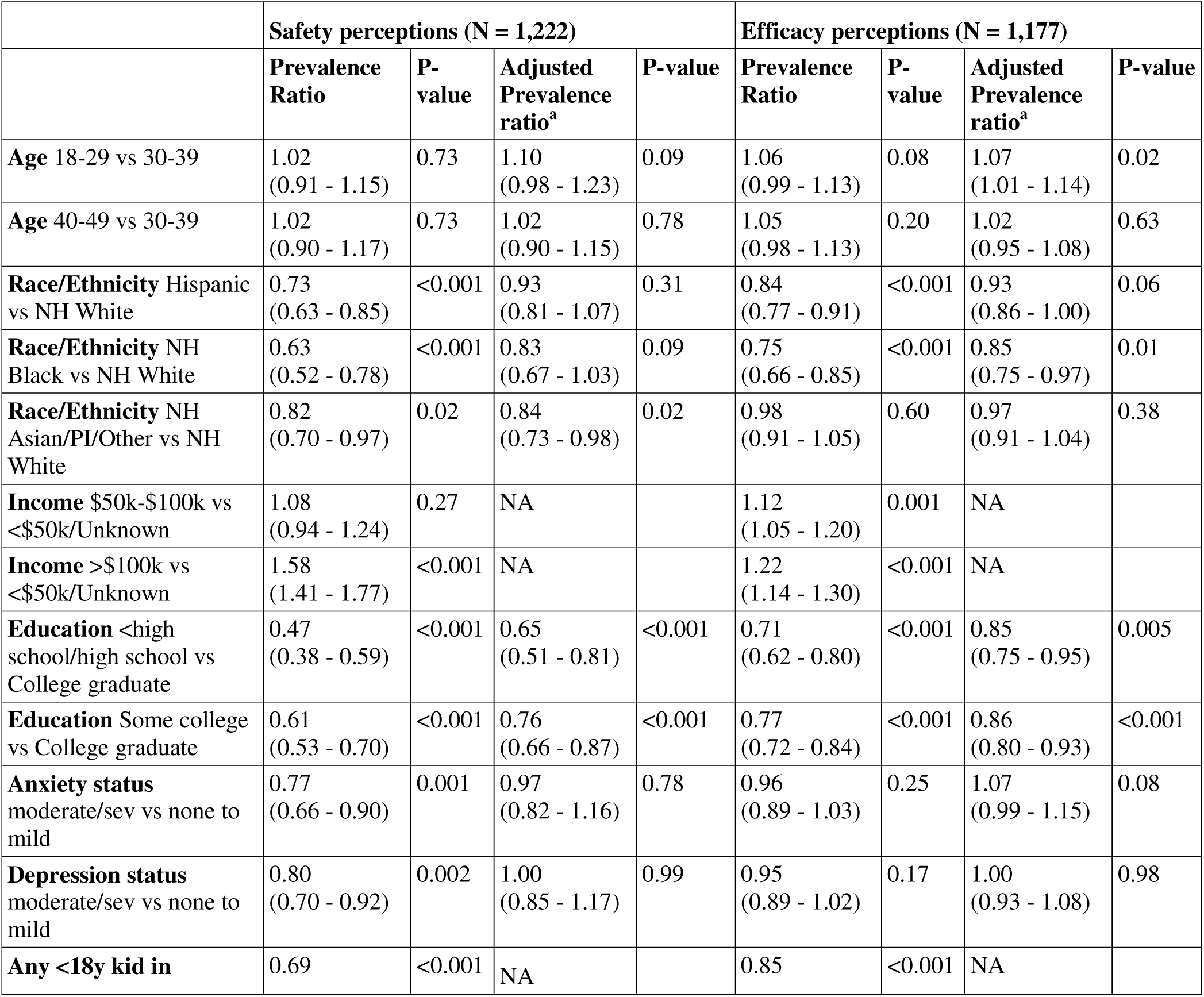
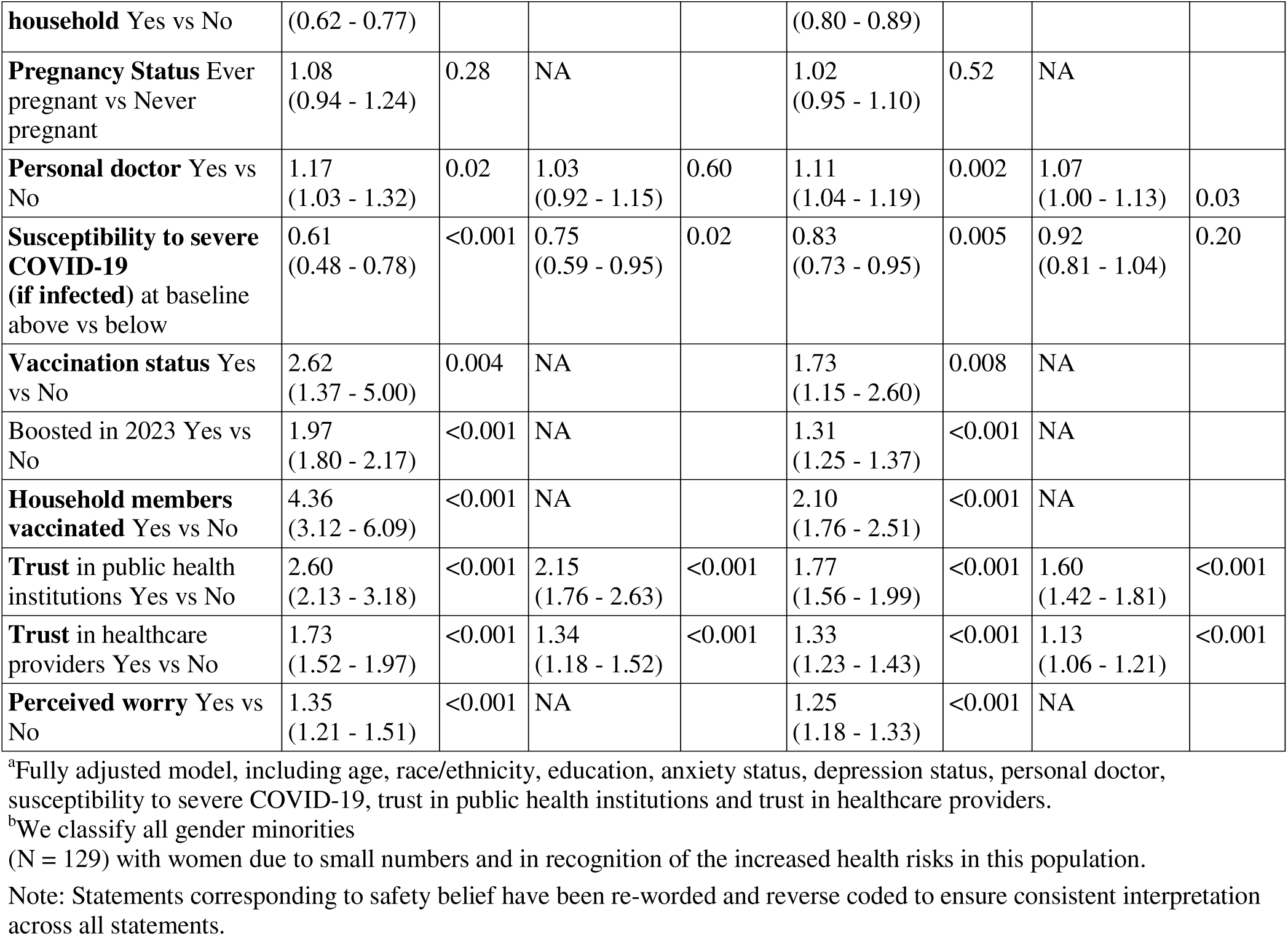
Prevalence Ratios and P-values for Any Agreement vs. Some Disagreement about COVID-19 Vaccine During Pregnancy Among Women^b^ of Reproductive Age in the CHASING COVID Cohort.

Any agreement with statements related to the efficacy of the COVID-19 vaccine during pregnancy was positively associated with trust in healthcare providers (aPR=1.13, 95% CI: 1.06–1.21), trust in public health institutions (aPR=1.60, 95% CI: 1.42–1.81), and having a personal doctor (aPR=1.07, 95% CI: 1.00–1.13).

#### Any agreement vs Uncertainty model

Among all adults (S1 Table), having a personal doctor and trust in healthcare providers were significantly associated with agreement on the efficacy of the COVID-19 vaccine during pregnancy. Trust in public health institutions was strongly associated with both safety and efficacy perceptions, but it was the only significant predictor for safety-related statements. Among women of reproductive age (S2 Table), having a personal doctor was significantly associated with agreement on both the safety and efficacy of the COVID-19 vaccine during pregnancy. While trust in public health institutions and healthcare providers was significant for efficacy perceptions, they were not significant predictors for safety perceptions in this subgroup.

## 4. Conclusion

This study revealed that less than half of adults (40%) perceived the COVID-19 vaccine as safe during pregnancy, with just over half considering it effective. Perceptions were influenced by demographics, with middle-aged, non-Hispanic White individuals, women, and those with a college education more likely to view the vaccine as safe and effective. Low uptake reasons often cited fears of maternal sickness or harm, insufficient research, and potential harm to the fetus[19], or broader concerns about side effects, infertility, and mortality[20]. Conversely, having vaccinated family/friends or prior Tdap/influenza vaccination was associated with less hesitancy[21].

A key finding was the strong association between trust and positive vaccine perceptions. Individuals who trusted healthcare providers were significantly more likely to regard the vaccine as safe (1.44 times) and effective (1.24 times) during pregnancy. Similarly, trust in public health institutions for vaccine information correlated with perceiving the vaccine as safe (almost three times more likely) and effective (almost two times more likely). These results underscore the vital role of healthcare providers and governmental risk communicators in building trust.

Limitations include a restricted number of pregnant individuals, preventing assumptions of causality between beliefs and actual vaccine uptake. Women of reproductive age were thus included to address unique immunization concerns during/after pregnancy as they may become pregnant. Future research should further explore the relationship between vaccine beliefs and actual vaccination behavior during pregnancy, and consider the influence of various healthcare providers.

In conclusion, this study highlights the complex interplay of demographic factors, healthcare experiences, and trust in shaping perceptions of COVID-19 vaccination during pregnancy. These findings are consistent with prior literature that highlights the significant influence of physician recommendations and trust in healthcare providers on vaccination decisions during pregnancy [23,24], including both general providers and obstetricians/gynecologists[25]. Given suboptimal vaccination rates, current strategies may be insufficient. Efforts to improve vaccine uptake must prioritize fostering trust, both at the individual level through provider-patient relationships and at the institutional level via transparent, community-centered communication strategies.

## Data Availability

Data cannot be shared publicly because of ethical restrictions regarding participant privacy and the sensitive nature of the health data collected.

## Acknowledgments

Support for this project was provided by the CUNY Institute for Implementation Science in Population Health and the CUNY Graduate School of Public Health and Health Policy.

Support for the CHASING COVID Cohort Study was provided by the National Institute of Allergy and Infectious Diseases (NIAID), award number UH3AI133675 (MPIs: D Nash and C Grov), NIMH award RF1MH132360 (MPIs: D Nash and A Parcesepe), National Institute of Child Health and Human Development grant P2CHD050924 (Carolina Population Center), Pfizer Inc., the CUNY Institute for Implementation Science in Population Health (cunyisph.org), and the COVID-19 Grant Program of the CUNY Graduate School of Public Health and Health Policy. The funders played no role in the production of this manuscript nor necessarily endorse the findings.

The City University of New York (CUNY) School of Public Health received research support from Pfizer during the conduct of this study that was paid directly to CUNY, with Dr. Nash serving as the Principal Investigator. All authors declare no conflicts of interest related to this work.

## Supporting information

**S1 Table. Prevalence Ratios and P-values for Any Agreement vs. Uncertainty about COVID-19 Vaccine During Pregnancy in the CHASING COVID Cohort among all adult participants**

**S2 Table. Prevalence Ratios and P-values for Any Agreement vs. Uncertainty about COVID-19 Vaccine During Pregnancy in the CHASING COVID Cohort among all adult participants**

